# A Neanderthal OAS1 isoform Protects Against COVID-19 Susceptibility and Severity: Results from Mendelian Randomization and Case-Control Studies

**DOI:** 10.1101/2020.10.13.20212092

**Authors:** Sirui Zhou, Guillaume Butler-Laporte, Tomoko Nakanishi, David Morrison, Jonathan Afilalo, Marc Afilalo, Laetitia Laurent, Maik Pietzner, Nicola Kerrison, Kaiqiong Zhao, Elsa Brunet-Ratnasingham, Danielle Henry, Nofar Kimchi, Zaman Afrasiabi, Nardin Rezk, Meriem Bouab, Louis Petitjean, Charlotte Guzman, Xiaoqing Xue, Chris Tselios, Branka Vulesevic, Olumide Adeleye, Tala Abdullah, Noor Almamlouk, Yiheng Chen, Michaël Chassé, Madeleine Durand, Michael Pollak, Clare Paterson, Hugo Zeberg, Johan Normark, Robert Frithiof, Miklós Lipcsey, Michael Hultström, Celia M T Greenwood, Claudia Langenberg, Elin Thysell, Vincent Mooser, Vincenzo Forgetta, Daniel E. Kaufmann, J Brent Richards

**Author notes:** These authors contributed equally to this study (Sirui Zhou, Guillaume Butler-Laporte, Tomoko Nakanishi, J Brent Richards). **Corresponding author:** Brent Richards, Professor of Medicine, McGill University, Senior Lecturer, King’s College London (Honorary), Pavilion H-413, Jewish General Hospital, 3755 Côte-Ste-Catherine Montréal, Québec, Canada, H3T 1E2, E www.mcgill.ca/genepi. **Disclosures:** JBR has served as an advisor to GlaxoSmithKline and Deerfield Capital.

## Abstract

Proteins detectable in peripheral blood may influence COVID-19 susceptibility or severity. However, understanding which circulating proteins are etiologically involved is difficult because their levels may be influenced by COVID-19 itself and are also subject to confounding factors. To identify circulating proteins influencing COVID-19 susceptibility and severity we undertook a large-scale two-sample Mendelian randomization (MR) study, since this study design can rapidly scan hundreds of circulating proteins and reduces bias due to reverse causation and confounding. We identified genetic determinants of 931 circulating proteins in 28,461 SARS-CoV-2 uninfected individuals, retaining only single nucleotide polymorphism near the gene encoding the circulating protein. We found that a standard deviation increase in OAS1 levels was associated with reduced COVID-19 death or ventilation (N = 4,336 cases / 623,902 controls; OR = 0.54, P = 7×10^−8^), COVID-19 hospitalization (N = 6,406 / 902,088; OR = 0.61, P = 8×10^−8^) and COVID-19 susceptibility (N = 14,134 / 1,284,876; OR = 0.78, P = 8×10^−6^). Results were consistent in multiple sensitivity analyses. We then measured OAS1 levels in 504 patients with repeated plasma samples (N=1039) with different COVID-19 outcomes and found that increased OAS1 levels in a non-infectious state were associated with protection against very severe COVID-19, hospitalization and susceptibility. Further analyses suggested that a Neanderthal isoform of OAS1 affords this protection. Thus, evidence from MR and a case-control study supported a protective role for OAS1 in COVID-19 outcomes. Available medicines, such as phosphodiesterase-12 inhibitors, increase OAS1 and could be explored for their effect on COVID-19 susceptibility and severity.

## Introduction

To date, the COVID-19 pandemic has caused more than 1.6 million deaths worldwide, and infected over 75 million individuals.^1^ Despite the scale of the epidemic, there are at present few disease-specific therapies^2^. to reduce the morbidity and mortality of SARS-CoV-2 infection, and apart from dexamethasone therapy in oxygen dependent patients^3^, most clinical trials have shown at most mild or inconsistent benefits in disease outcome.^4–6^ Therefore, validated targets are needed for COVID-19 therapeutic development.

One source of such targets is circulating proteins. Recent advances in large-scale proteomics have enabled the measurement of thousands of circulating proteins at once and when combined with evidence from human genetics, such targets greatly improve the probability of drug development success.^7–9^ While *de novo* drug development will take time—even in the accelerated arena of COVID-19 therapies— repurposing of currently available molecules targeting those proteins could also provide an accelerated opportunity to deliver new therapies to patients.

Nevertheless, since confounding and reverse causation often bias traditional circulating protein epidemiological studies, disentangling the causal relationship between circulating proteins and COVID-19 susceptibility or severity is challenging. This is especially the case in COVID-19, where exposure to SARS-CoV-2 unleashes profound changes in circulating protein levels^10^. One way to address these limitations is by using Mendelian randomization (MR), a genetic epidemiology method that uses genetic variants as instrumental variables to test the effect of an exposure (here protein levels) on an outcome (here COVID-19 outcomes). Given that genotypes are determined by randomly segregated alleles during meiosis of parental gametes, this greatly reduces bias due to confounding. Since genotypes are always assigned prior to disease onset, MR studies are not influenced by reverse causation. However, MR rests on several assumptions^11^, the most problematic being the lack of horizontal pleiotropy of the genetic instruments (wherein the genotype influences the outcome, independently of the exposure). One way to help avoid this bias is to use genetic variants that influence circulating protein levels which are adjacent to the gene which encodes the circulating protein through the use of *cis*-protein quantitative trait loci (*cis*-pQTLs).^9^ Given their close proximity to the target gene, *cis*-pQTLs are likely to influence the level of the circulating protein, among others, by directly influencing its transcription or translation, and therefore less likely to affect the outcome of interest (COVID-19) through pleiotropic pathways. Nevertheless, a causal genetic association between the exposure and outcome may be confounded by linkage disequilibrium (LD, the non-random association of genetic variants assigned at conception).^12^ To probe this potential problem, colocalization tests can assess for the presence of bias from LD.

Understanding the etiologic role of circulating proteins in infectious diseases is challenging because the infection itself often leads to large changes in circulating protein levels^10^. Thus, it may appear that an increase in a circulating protein, such as a cytokine, is associated with a worsened outcome, when in fact, the cytokine may be the host’s response to this infection and help to mitigate this outcome. It is therefore important to identify genetic determinants of the protein levels in the non-infected state, which would reflect a person’s baseline predisposition to the level of a protein.

MR studies can be complemented by traditional case-control studies, where the protein is longitudinally measured in COVID-19 patients and controls, allowing for an estimation of the association between the protein level and COVID-19 outcomes. However, MR studies would tend to predict the effect of the protein in the non-infectious state when the genetic determinants of such proteins are measured in the non-infected population. Thus, longitudinal measurements of proteins can allow for a better understanding of the role of such proteins in COVID-19 outcomes and also describe how their levels respond to the infection. Since MR and case-control studies rely on different assumptions, and may be influenced by different biases, concordant results between the two study designs can strengthen the cumulative evidence through the concept of the triangulation of evidence^13^.

In this study, we therefore undertook two-sample MR and colocalization analyses to combine results from large-scale genome-wide association studies (GWAS) of circulating protein levels and COVID-19 outcomes^14^ in order to prioritize proteins likely influencing COVID-19 outcomes. We began by identifying the genetic determinants of circulating protein levels in large-scale protein level GWASs, then used MR to assess whether these *cis*-pQTLs were associated with COVID-19 outcomes in the ICDA Host Genetics Initiative COVID-19 outcomes GWASs. Next, we investigated expression QTL (eQTL) and splice QTL (sQTL) effects of our lead proteins. We then measured the most promising protein, OAS1, in 504 subjects ascertained for SARS-CoV-2 infection and when PCR positive, followed for longitudinal sampling during and after their infection.

## Results

### MR using cis-pQTLs, and pleiotropy assessment

Study design is illustrated in **Figure 1**. We began by obtaining the genetic determinants of circulating protein levels from six large proteomic GWAS of European individuals (Sun *et al*^15^ N=3,301; Emilsson *et al*^16^ N=3,200; Pietzner *et al*^17^ N=10,708; Folkersen *et al*^18^ N=3,394; Yao *et al*^19^ N=6,861 and Suhre *et al*^20^ N=997). A total of 931 proteins from these six studies had *cis*-pQTLs associated at a genome-wide significant level (P < 5×10^−8^) with protein levels, or highly correlated proxies (LD R^2^ > 0.8), in the meta-analyses of data the from COVID-19 Host Genetics Initiative^21^ which included results from the GenOMICC program^22^. We then undertook MR analyses using 1,425 directly matched *cis-*pQTLs and 39 proxies as genetic instruments across six studies for their associated circulating proteins on three separate COVID-19 outcomes: 1) Very severe COVID-19 disease (defined as individuals experiencing death, mechanical ventilation, non-invasive ventilation, high-flow oxygen, or use of extracorporeal membrane oxygenation. 99.7% of these individuals were of European ancestry) using 4,336 cases and 623,902 controls; 2) COVID-19 disease requiring hospitalization using 6,406 cases and 902,088 controls of European ancestry and 3) COVID-19 susceptibility using 14,134 cases and 1,284,876 controls of European ancestry. These case-control phenotype definitions are referred to as A2, B2, and C2 by the COVID-19 Host Genetics Initiative, respectively. In all outcomes, cases required evidence of SARS-CoV-2 infection. For the very severe COVID-19 and hospitalization outcomes, COVID-19 cases were defined as laboratory confirmed SARS-CoV-2 infection based on nucleic acid amplification or serology tests. For the COVID-19 susceptibility outcome, cases were also identified by review of health records (using International Classification of Disease codes or physician notes).

**Figure 1.**
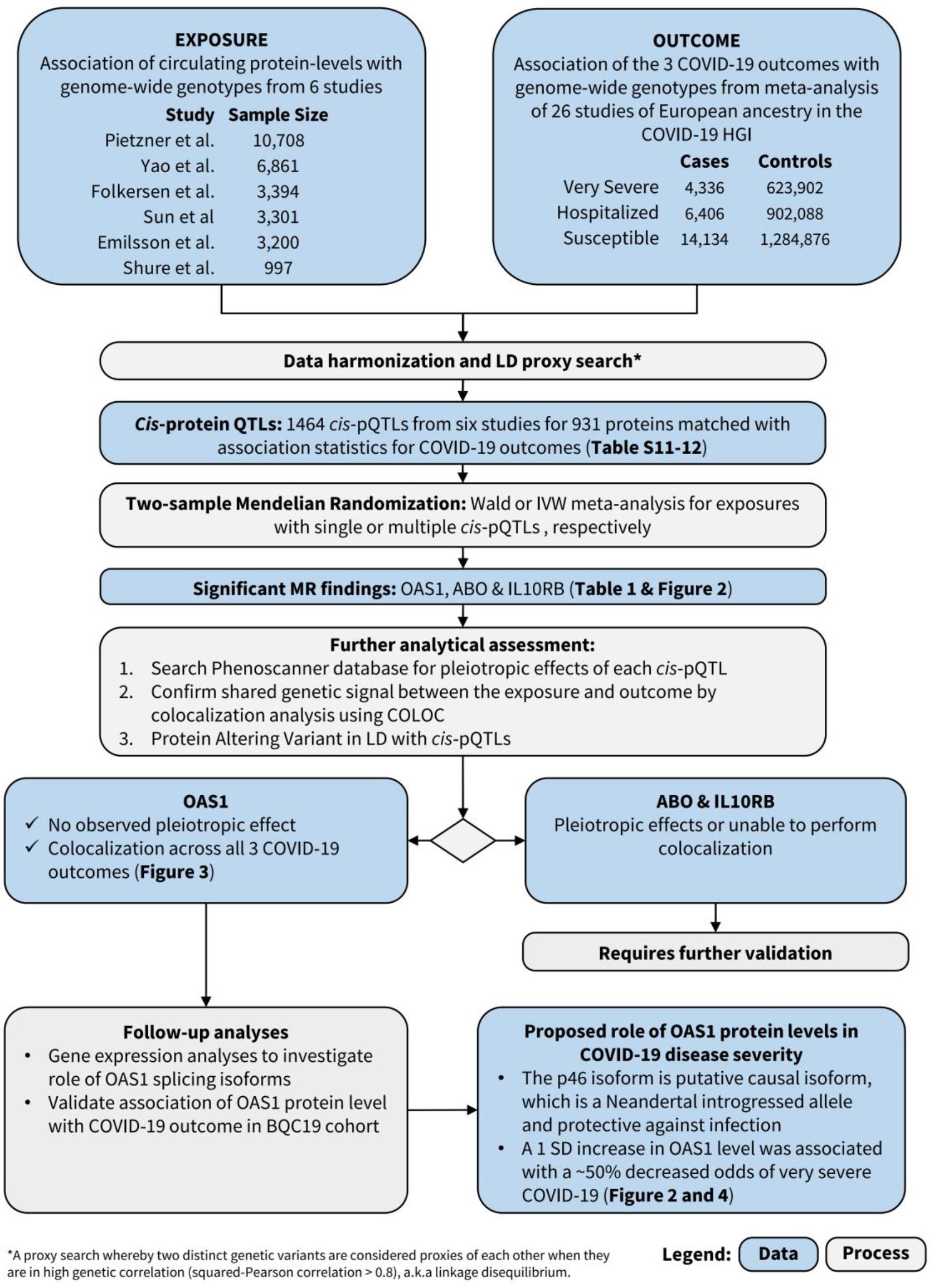
Flow Diagram of Study Design.

MR analyses revealed that the levels of three circulating proteins, 2’-5’-oligoadenylate synthetase 1 (OAS1), interleukin-10 receptor beta subunit (IL10RB) and ABO were associated with at least two COVID-19 outcomes after Benjamini & Hochberg FDR correction for the number of proteins tested (**Table 1, Tables S1-6**). We note that FDR correction is overly conservative given the non-independence of the circulating protein levels. Notably, increased OAS1 levels were strongly associated with protection from all three COVID-19 outcomes. Further, these effect sizes were more pronounced in severe and hospitalization outcomes, such that each standard deviation increase in OAS1 levels was associated with decreased odds of very severe COVID-19 (OR=0.54; 95% CI: 0.44-0.68, P=7.0×10^−8^), hospitalization (OR=0.61; 95% CI: 0.51-0.73, P=8.3×10^−8^) and susceptibility (OR=0.78; 95% CI: 0.69-0.87, P=7.6×10^−6^) (**Figure 2A**). We also identified OAS1 *cis*-pQTLs in Emilsson *et al*^16^ and Pietzner *et al*^17^ which were not included in the MR analyses due to lack of genome-wide significance in their association with OAS1 levels or missing from initial protein panel. Undertaking MR analyses of using these additional *cis*-pQTLs, we found concordant results (**Table S7**).

**Table 1.** MR-Identified Circulating Protein Levels Effecting COVID-19 Outcomes.

| Protein | *cis*-pQTL | Source | Very Severe COVID-19<br>(99.7% European Ancestry) |  |  |  | Hospitalization<br>(European Ancestry Only) |  |  |  | Susceptibility<br>(European Ancestry Only) |  |  |  |
| --- | --- | --- | --- | --- | --- | --- | --- | --- | --- | --- | --- | --- | --- | --- |
|  |  |  | OR | 95%CI | P value | P het | OR | 95%CI | P value | P het | OR | 95%CI | P value | P het |
| OAS1 | rs4767027 | Sun | 0.54 | 0.44-0.68 | $7.0 \times 10^{-8}$ | 0.37 | 0.61 | 0.51-0.73 | $8.3 \times 10^{-8}$ | 0.16 | 0.78 | 0.69-0.87 | $7.6 \times 10^{-6}$ | 0.005 |
| ABO | rs505922 | Sun, Emilsson | 1.09 | 1.05-1.14 | $6.4 \times 10^{-5}$ | 0.10 | 1.11 | 1.07-1.15 | $6.8 \times 10^{-9}$ | 0.06 | 1.07 | 1.05-1.10 | $1.1 \times 10^{-9}$ | 0.10 |
| IL10RB | rs2834167 | Emilsson | 0.47 | 0.32-0.68 | $7.1 \times 10^{-5}$ | 0.02 | 0.53 | 0.39-0.73 | $8.8 \times 10^{-5}$ | 0.11 | 0.87 | 0.72-1.07 | 0.18 | 0.006 |

OR: represents the estimated effect of a standard deviation on the natural log scale (for Sun et al) or one unit (for Emilsson et al) increase in protein levels on the odds of the three COVID-19 outcomes. P het: P value of heterogeneity for each *cis*-pQTLs across the cohorts in the GWAS summary-level meta-analysis from COVID-19 Host Genomic Initiative.

**Figure 2.**
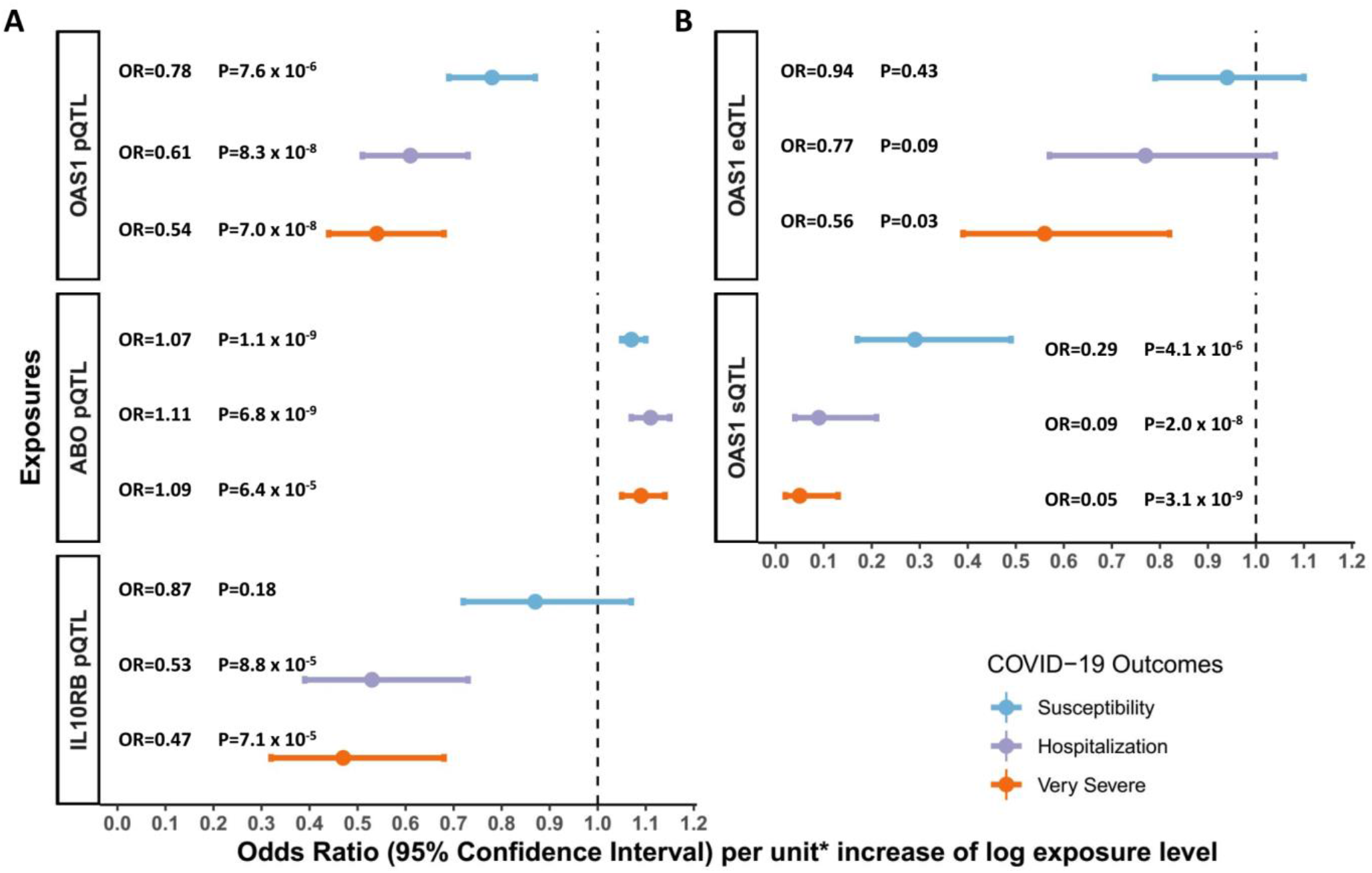
Association of Circulating Protein Levels of OAS1, ABO and IL10RB and mRNA levels of OAS1 with COVID-19 Outcomes from MR. A: MR estimates of proteins influencing COVID-19 outcomes, unit: standard deviation of log normalized value; B. MR estimates of OAS1 mRNA influencing COVID-19 outcomes, unit: standard deviation of normalized read counts.

We next assessed whether the *cis-*pQTL associated with OAS1 levels (rs4767027) was associated with any other phenotypes across more than 5,000 outcomes, as catalogued in PhenoScanner,^23^ which collects associations of SNPs with outcomes from all available GWASs. We found that the only significant association for rs4767027 was with circulating OAS1 levels (P=6.2×10^−26^) in plasma, whereas it was not associated with any other traits or protein levels (P<5.0×10^−5^). These findings reduce the possibility that the MR estimate of the effect of OAS1 on COVID-19 outcomes is due to horizontal pleiotropy. Finally, except for the susceptibility outcome, the effect of rs4767027 did not demonstrate evidence of heterogeneity across COVID-19 Host Genetics Initiative GWAS meta-analyses (**Table 1)**.

We next identified an independent SNP associated with OAS1 circulating protein levels, which was not at the *OAS1* locus and is thus a *trans*-SNP (rs62143197, P value for association with OAS1 levels =7.10 x 10^−21^). However, this SNP is likely subject to pleiotropic effects, since it is strongly associated with many other proteins, such as annexin A2 (P=5.6 x 10^−237^) and small ubiquitin-related modifier 3 (P=9.1 x 10^−178^). Consequently, including this *trans*-SNP could introduce bias from horizontal pleiotropic effects and was thus not considered in further MR analyses. Further, this trans-association signal was unique to the INTERVAL study^17^.

OAS proteins are part of the innate immune response against RNA viruses. They are induced by interferons and activate latent RNase L, resulting in direct viral and endogenous RNA destruction, as demonstrated in *in-vitro* studies.^24^ Thus OAS1 has a plausible biological activity against SARS-CoV-2.

Using a *cis-*pQTL for IL10RB (rs2834167), we found that one standard deviation increase in circulating IL10RB level was associated with decreased odds for very severe COVID-19 (OR=0.47; 95% CI: 0.32-0.68, P=7.1×10^−5^) and hospitalization (OR = 0.53; 95% CI: 0.39-0.73, P=8.8×10^−5^). However, circulating IL10RB protein level was not associated with COVID-19 susceptibility. Using PhenoScanner, we could not find evidence of pleiotropic effects of the *cis-*pQTL for IL10RB. The IL10RB *cis-*pQTL also showed a homogeneous effect across the three COVID-19 outcomes except for susceptibility to COVID-19 (**Table 1, Figure 2A**). MR revealed that one standard deviation increase in circulating ABO level was associated with increased odds of adverse COVID-19 outcomes (**Table 1**), however, we found that a *cis-*pQTL for ABO (rs505922) was strongly associated with the levels of several other proteins, suggesting potential horizontal pleiotropic effects (**Table S8**). Given ABO’s known involvement in multiple physiological processes, these results were expected, but highlight that MR analyses may suffer from significant bias from horizontal pleiotropy.

### Colocalization Studies

To test whether confounding due to LD may have influenced the estimated effect of circulating OAS1 on the three different COVID-19 outcomes, we tested the probability that the genetic determinants of OAS1 circulating protein level were shared with the three COVID-19 outcomes using colocalization analyses. These were performed using *coloc*, a Bayesian statistical test implemented in the *coloc* R package.^12^ We found that the posterior probability that OAS1 levels and COVID-19 outcomes shared a single causal signal (the posterior probability for hypothesis 4 in *coloc*, PP4) in the 1Mb locus around the cis-pQTL rs4767027 was 0.72 for very severe COVID-19, 0.82 for hospitalization due to COVID-19, and 0.89 for COVID-19 susceptibility (**Figure 3)**. This colocalization result was also replicated using *cis*-pQTLs for OAS1 levels identified by Pietzner *et al*^17^ (**Table S7**). This suggests that there is likely a single shared causal signal for OAS1 circulating protein levels and COVID-19 outcomes.

**Figure 3.**
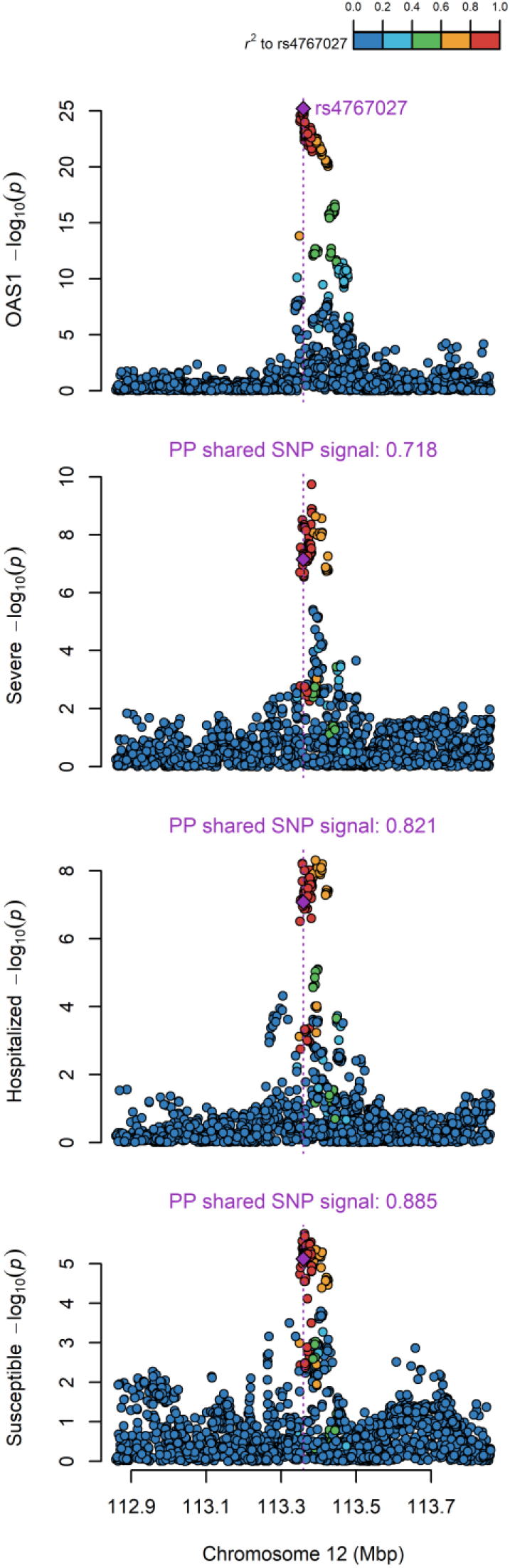
Colocalization of the Genetic Determinants of OAS1 Plasma Protein Levels and COVID-19 Outcomes. Colocalizatinon of genetic signal of 1MB region around OAS1 pQTL rs4767027 of OAS1 level (top plot) and COVID-19 outcomes (three bottom plot), color shows SNPs in the region in LD (r^2^) to rs4767027 (purple). Posterial probability (PP) of shared signal between OAS1 level and three COVID-19 ouctomes are estimated by *coloc*.

Colocalization of ABO levels and different COVID-19 outcomes also showed colocalization between ABO level and different COVID-19 outcomes (posterior probability of single shared signal = 0.90, 0.98 and 1 for ABO level and very severe COVID-19, hospitalization due to COVID-19 and susceptibility, respectively) (**Figure S1)**. We were unable to perform colocalization analyses for IL10RB due to a lack of genome-wide summary level data from the original proteomic GWAS^16^.

### Aptamer Binding Effects

Protein altering variants (PAVs)^15^ may influence binding of affinity agents, such as aptamers or antibodies, that are used to quantify protein levels. We thus assessed if the *cis-*pQTLs for the MR-prioritized proteins were PAVs, or in LD (R^2^>0.8) with PAVs, and if so, whether conditioning the *cis*-pQTLs on correlated PAVs influenced their association with COVID-19 outcomes. rs2834167 (IL10RB) is a nonsense variant and could therefore be subject to potential binding effects. rs505922 (ABO) is not in LD with known missense variants. rs4767027 (OAS1) is an intronic variant, which is in LD with a missense variant rs2660 (R^2^=1) in European ancestry. Unfortunately, this missense variant was not included in the imputation data of Sun *et al*, and the effect by this missense variant could not be evaluated. However, since RNA splicing and expression studies derived from RNA sequencing are not subject to potential effects of missense variants that could influence aptamer binding, we next explored whether rs4767027 also influences OAS1 splicing and/or expression.

### sQTL and eQTL studies for OAS genes

Splicing QTLs (sQTLs) are genetic variants that influence the transcription of different isoforms of a protein. The aptamer that targets OAS1 was developed against a synthetic protein comprising the amino acid sequence 1-364 of NP002525.2^25^, which is common to the two major OAS1 isoforms, p46 and p42, and hence the aptamer may identify both, or either isoforms. rs10774671 is a known sQTL for OAS1 that induces alternate splicing and create p46 and p42, a majority of present-day people carry this splice variant (rs10774671-A), which increases expression of isoforms other than p46^26^. The ancestral variant (rs10774671-G) is the major allele in African populations and became fixed in Neanderthal and Denisovan genomes^27,28^. However, the ancestral variant, with its increased expression of the p46 isoform, was reintroduced into the European population via gene flow from Neanderthals^29^, and is also the predominant isoform found in circulating blood^26^. The p46 isoform has been demonstrated to have higher anti-viral activity than other isoforms^30^. Interestingly, the OAS1 pQTL, rs4767027, is in high LD (R^2^=0.97) with rs10774671^29^ in European populations. Functional studies support that the G allele at rs10774671 increases expression of the p46 isoform but decreases expression of the p42 isoform^26^. This G allele at the sQTL rs10774671 reflects the T allele at pQTL rs4767027, which itself is associated with higher measured OAS1 levels and reduced odds of COVID-19 severity and susceptibility. These separate lines of evidence suggest that the p46 isoform was predominantly measured by the SomaScan^®^ platform and may protect against COVID-19 outcomes.

Undertaking MR studies of OAS1 splicing, we found that increased expression of the p46 isoform (as defined by normalized read counts of the intron cluster defined by LeafCutter^31,32^) was associated with reduced odds of COVID-19 outcomes (OR = 0.29; 95% CI: 0.17-0.49, P=4.1×10^−6^ for susceptibility, OR = 0.09; 95% CI: 0.04-0.21, P=2.0×10^−8^ for hospitalization and OR = 0.05; 95% CI: 0.02-0.13, P=3.1×10^−9^ for very severe COVID-19) (**Figure 2B**). Colocalization analyses also supported a shared causal signal between the sQTL for *OAS1*, the pQTL and COVID-19 outcomes (**Figure S2A-B**). Interestingly, the colocalization analyses supported a stronger probability of a shared signal with the sQTL, than the pQTL, suggesting that the p46 isoform may be the driver of the association of OAS1 levels with COVID-19 outcomes.

Next, we tested whether increased expression of OAS1 levels, without respect to isoform, were associated with COVID-19 outcomes using eQTL MR analyses. We identified an expression QTL (eQTL) for total OAS1, rs10744785, from GTEx v8.^33^ Total *OAS1* expression levels were not associated with COVID-19 susceptibility and hospitalization (**Figure 2B**). We also found that increased *OAS3* expression level in whole blood was positively associated with COVID-19 outcomes in MR analyses with a support for colocalization of their genetic signal (**Table S9, Figure S3**).

Taken together, these pQTL, sQTL and eQTL studies suggest that increased levels of the p46 isoform of OAS1 protect against COVID-19 adverse outcomes. Further, the concordant evidence from sQTL and pQTL MR studies suggest that the effect of OAS1 levels on COVID-19 outcomes is unlikely to be biased by aptamer binding effects.

### Association of measured OAS1 protein level with COVID-19 outcomes

Since MR studies were derived from protein levels measured in a non-infected state, we tested the hypothesis that increased OAS1 protein levels in a non-infected state would be associated with reduced odds of COVID-19 outcomes. To do so, we undertook a case-control study, measuring OAS1 protein levels using the SomaScan^®^ platform in 1039 longitudinal samples from 399 SARS-CoV-2 PCR positive patients collected at multiple time points during their COVID-19 infection and 105 individuals who presented with COVID-19 symptoms but had negative SARS-CoV-2 PCR nasal swabs from the Biobanque Quebecoise de la COVID-19 cohort (www.BQC19.ca). Individuals were recruited prospectively who had undergone nasal swabs for SARS-CoV-2 infection. The demographic characteristics of the participants in the BQC19 cohort who underwent SomaScan^®^ assays is detailed in **Table 2**.

**Table 2.**
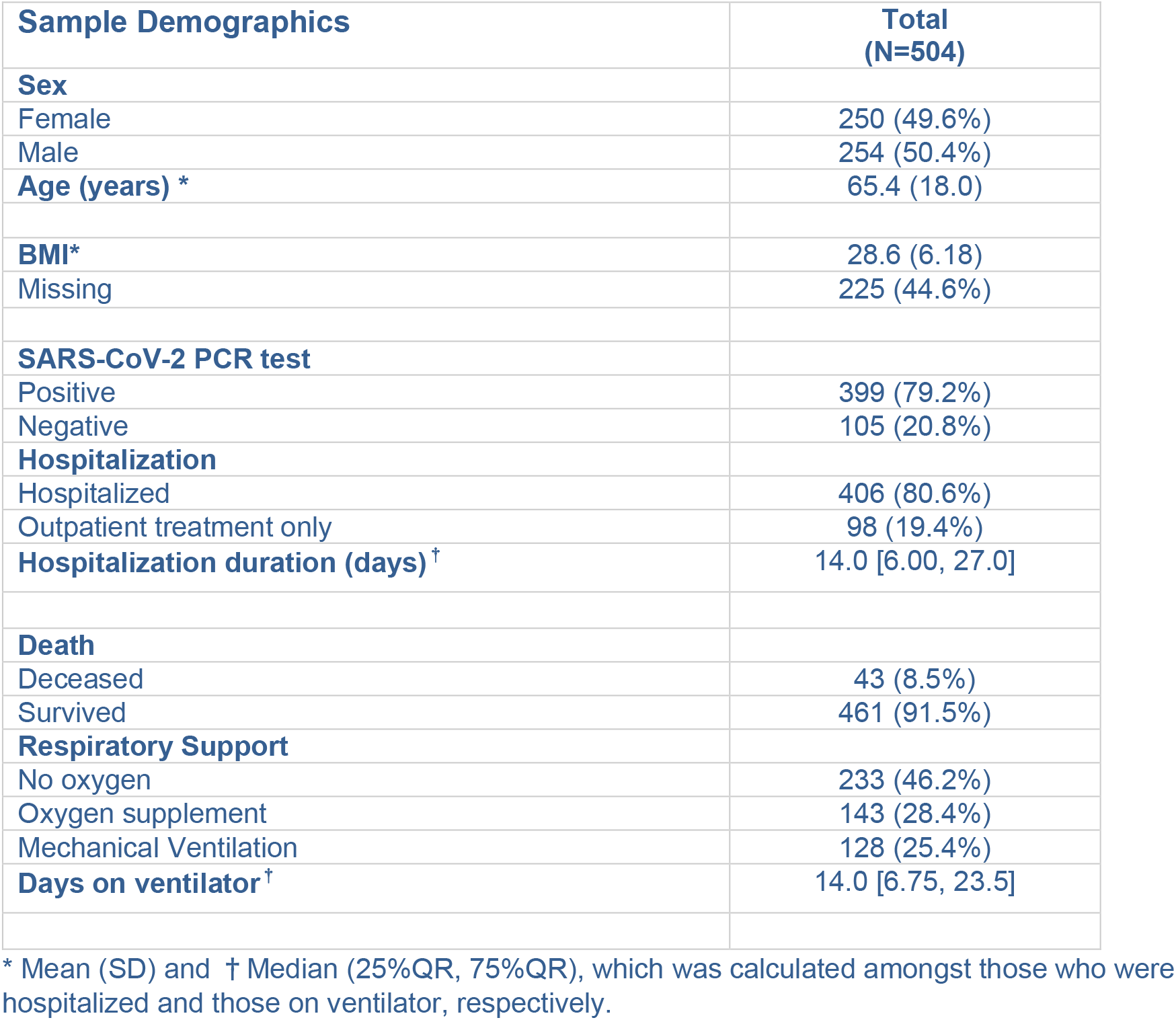
Participant demographics for the BQC19 cohort.

We defined non-infectious samples as those collected from convalescent SARS-CoV-2 patients at least 31 days after onset of their symptoms (N=115), or samples collected from SARS-CoV-2 PCR negative patients (N=105). These SARS-CoV-2 PCR negative patients were recruited as controls into the study, as their inclusion reduces the probability of the introduction of collider bias^34^. As we also observed a change in OAS1 level with the exposure to the SARS-CoV-2 virus (**Figure S4**), in order to understand how OAS1 protein levels during infection would be associated with COVID-19 outcomes, we also measured OAS1 levels in individuals with samples from SARS-CoV-2 positive patients <14 days after symptom onset (N=313). Sample outliers were removed (**Figure S5, S6**), and we showed that OAS1 levels are not associated with age and sex in samples without active infection (**Figure S7**). Additional sample QC and characterization of the cohort is described in **Supplementary data**.

To test whether OAS1 levels in a non-infectious state were associated with COVID-19 outcomes we undertook logistic regression using the three COVID-19 outcomes, while controlling for age, sex, age^2^, plate, recruitment center and sample processing time. OAS1 levels were log-transformed and standardized to match the transformation procedure of the MR study. We found that in the non-infectious samples, each standard deviation increase in OAS1 levels on the log-transformed scale was associated with reduced odds of COVID-19 outcomes (OR = 0.20 [95% CI: 0.08 – 0.53]; P = 0.001 for very severe COVID-19, OR = 0.46 [95% CI: 0.28 – 0.76], P = 0.002 for hospitalization and OR = 0.69 [95% CI: 0.49 – 0.98], P = 0.04 for susceptibility) (**Figure 4, Table S10, Figure S8**). These results are consistent with our findings from MR, where increased circulating OAS1 levels in a non-infectious state were associated with protection against all of these adverse COVID-19 outcomes.

**Figure 4.**
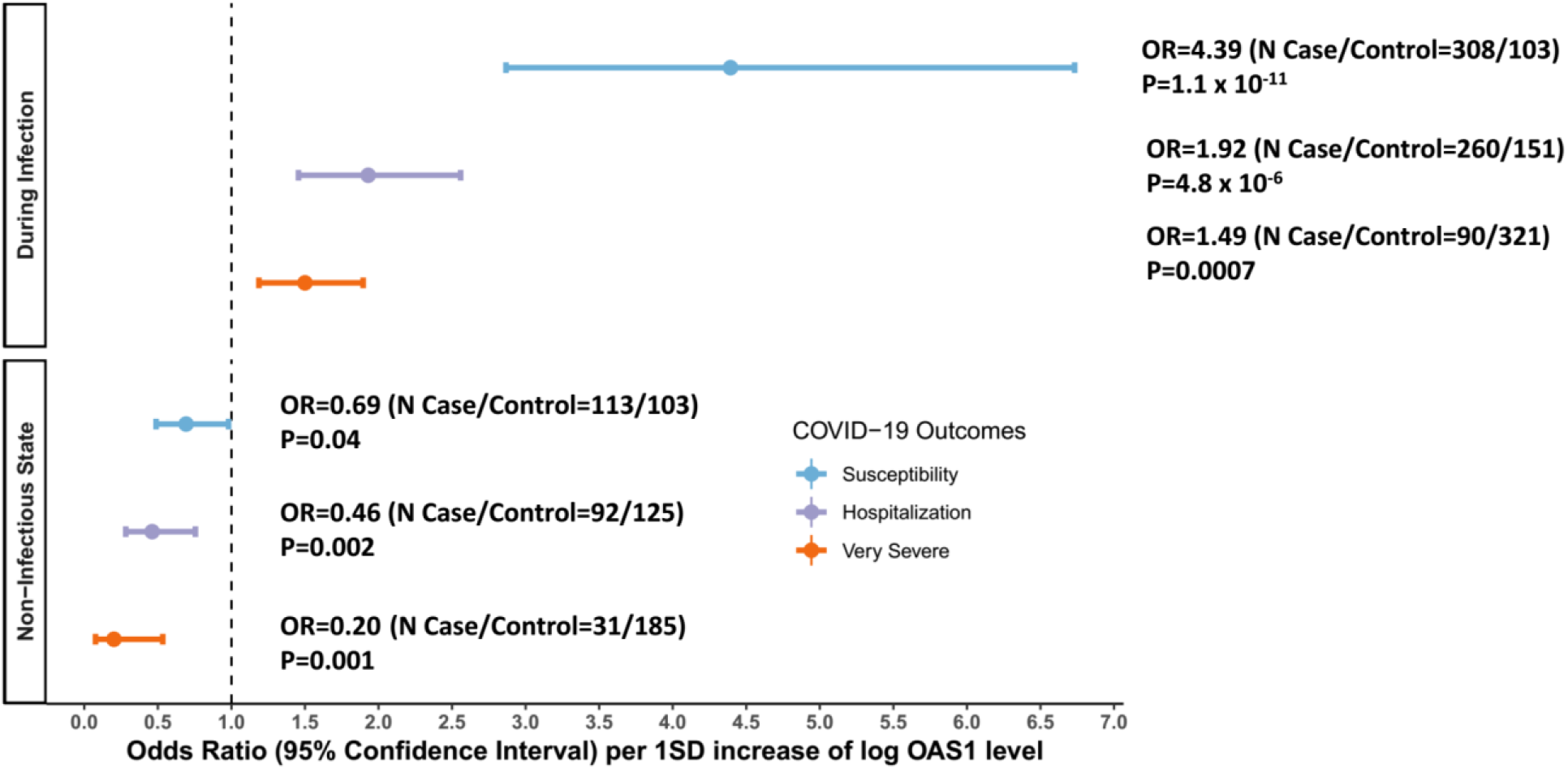
Association of OAS1 levels with COVID-19 Outcomes from the Case-Control Study in BQC19. During Infection: Patient samples that were collected within 14 days from the date of symptom onset. For individuals with two or more samples collected within 14 days of symptom onset, the earliest time point was used. Non-Infectious State: Patient samples that were collected at least 31 days from the date of symptom onset. For individuals with two or more samples collected at different time points at least 31 days from symptom onset, the latest time point was used. Additional information is also described in table S10.

In samples drawn during active infection we found that increased OAS1 levels were associated with increased odds of adverse COVID-19 outcomes (OR = 1.49 [95% CI: 1.19 – 1.90]; P = 0.0007 for very severe COVID-19, OR = 1.92 [95% CI: 1.46 – 2.56], P = 4.8 x 10^−6^ for hospitalization and OR = 4.39 [95% CI: 2.87 – 6.73], P = 1.09 x 10^−11^ for susceptibility) (**Figure 4, Table S10, Figure S8**).

Taken together, these findings suggest that increased OAS1 levels in a non-infectious state are associated with better COVID-19 outcomes, and that during infection, SARS-CoV-2 exposure likely causes OAS1 levels to increase, as interferon pathways are stimulated, which are known to increase OAS1 levels^35^.

## Discussion

Disease-specific therapies are needed to reduce the morbidity and mortality associated with COVID-19 outcomes. In this large-scale two-sample MR study of 931 proteins assessed for three COVID-19 outcomes in up to 14,134 cases and 1.2 million controls with European ancestry, we provide evidence that increased OAS1 levels in the non-infectious state are strongly associated with reduced risks of very severe COVID-19, hospitalization and susceptibility. The protective effect size was particularly large, such that a 50% decrease in the odds of very severe COVID-19 was observed per standard deviation increase in OAS1 circulating levels. Since therapies exist that activate OAS1, repositioning them as potential COVID-19 treatments should be prioritized.

In non-Sub-Saharan African populations, the protective alleles at both rs4767027-T (the OAS1 pQTL) and rs10774671-G (the OAS1 sQTL) are found on a Neandertal haplotype which was passed on to modern humans ∼50-60,000 years ago^36^. Even though these two SNPs share a haplotype, their evolutionary histories differ. The rs4767027-T allele is derived from the Neandertal lineage, whereas for the rs10774671-G allele, Neanderthals preserved the ancestral state. OAS1 alternative splicing regulated by the rs10774671-G allele increases the isoform p46, which is known to have a higher enzymatic activity against viruses than the p42 isoform^37^. p46 is also known to be the only OAS1 isoform which is robustly upregulated during infection^29^. Although further studies are needed to fully elucidate the functional relevance of the pQTL and sQTL for OAS1, the antiviral activity of the gene products is higher for the Neandertal haplotype than the common haplotype in Europeans^30^. In Europeans the Neandertal haplotype has undergone positive selection^29^ and the rs4767027-T allele reaches an allele frequency of 0.32, whereas it is absent in sub-Saharan African populations. The association between the Neanderthal haplotype and protection against severe COVID-19 was recently described^38^. Using MR and measurements of circulating proteins, we demonstrated here that increased OAS1 levels of the Neandertal haplotype confers this protective effect.

Our MR evidence indicated that higher p46 isoform levels of OAS1 and higher OAS1 total protein levels, as measured by the SomaScan^®^ assay had protective effects on COVID-19 outcomes. These results were strongly supported by colocalization analysis. Given the consistent colocalization between the sQTL and pQTL for OAS1, the lack of colocalization between the eQTL and pQTL for OAS1, and the evidence that the SomaScan^®^ assay likely measures p46 isoforms, rather than total protein levels, it seems probable that the protective effect of OAS1 is derived from the p46 isoform. However, further investigations are required to specifically measure each isoform in circulation.

In light of the protective effect of the ancestral OAS1 splice variant (rs10774671-G) on COVID-19 and the positive selection of the Neandertal haplotype in Europeans, the loss-of-function variant (rs10774671-A) found in non-African population is surprising. Several scenarios might explain this loss-of-function, e.g., loss of purifying selection during the out-Africa exodus due to changes in environmental pathogens. Moreover, immune responses can be harmful and loss-of-function in OAS1-antiviral activity has been observed in several primates^39^, suggesting a cost of OAS1 activity. Nevertheless, our results indicate that interbreeding between Neanderthals and modern humans confers some protection against COVID-19. The OAS1 Neanderthal variant is another risk-modulating locus reported to be inherited from Neanderthals, the other being the chromosome 3 risk locus^40^.

*OAS1, OAS2* and *OAS*3 share significant homology and differ only in their number of OAS units. They also increase expression of both IRF3 and IRF7, both genes involved in interferon-induced gene expression. As an interferon stimulated gene^41^, *OAS1* polymorphisms have been associated with the host immune response to several classes of viral infection including influenza^42^, herpes simplex^43^, hepatitis C, West Nile^44^ Dengue^45^, and SARS-CoV^46^ viruses. Given that OAS1 is an intracellular enzyme leading to viral RNA degradation, it is probable that the circulating levels of this enzyme reflect intracellular levels of this protein. However, there exists considerable evidence that circulating OAS1 is also important in the viral immune response^47^.

Molecules currently exist which can increase OAS1 activity. Interferon beta-1b, which activates a cytokine cascade leading to increased *OAS1* expression,^48^ is currently used to treat multiple sclerosis and has been shown to induce *OAS1* expression in blood.^49^ Interferon-based therapy has also been used in other viral infections^50^. However, recent randomized trials have shown inconsistent results. While intravenous interferon beta-1b combined with lopinavir-ritonavir reduced mortality due to MERS-CoV infections,^51^ in the unblinded SOLIDARITY trial,^52^ there was no demonstrated benefit of intravenous interferon-beta-1b. On the other hand, a recent phase II trial testing the effect of inhaled nebulized interferon beta-1b showed improved symptoms in the treatment arm.^53^ While this study was not powered to show a difference in mortality, all deaths occurred in the placebo group. Inhaled nebulized interferon-1-beta results in a much higher tissue availability in the lung and may result in improved anti-viral activity. Moreover, timing of administration is likely to play a role, as the administration of a pro-inflammatory cytokine may not provide benefit during the inflammation driven phase of the disease. However, data on timing of administration is currently unavailable in the SOLIDARITY trial, and conclusions cannot yet be drawn. Lastly the effect of interferon supplement may vary across ancestral population, as different ancestries have different amounts of the more active p46 isoform of OAS1. Our study was limited to individuals of European ancestry, a population with higher expression of the p46 isoform. Interestingly, the SOLIDARITY trial enrolled 61% of its patients in Africa or Asia, and 17% in Latin America, populations with higher expression of the p42 isoform OAS1, while the study on inhaled interferon beta-1b was comprised of 80% White patients from the United Kingdom. This suggests that interferon beta-1b may have different effects in populations of different ancestry, due to presence of different genetic variants.

*In-vitro* evidence also exists demonstrating that pharmacological inhibition of phosphodiesterase-12, which normally degrades the OAS enzymes, potentiates this OAS-mediated antiviral activity.^54^ PDE-12 inhibitors potentiate the action of OAS1, 2 and 3.^55^ Interestingly other coronaviruses in the same betacoronavirus family as SARS-CoV-2 have been shown to produce viral proteins that degrade the OAS family of proteins, and antagonize RNase-L activity, leading to evasion of the host immune response.^56,57^ Thus classes of medications currently exist that lead to increased OAS1 levels and could be explored for their effect upon COVID-19 outcomes.

Our MR analyses found that higher level of OAS3 expression is associated with worse COVID-19 outcomes, which is an opposite direction of effect compared to OAS1. The discordant effects of the Neanderthal haplotype for OAS1 and OAS3 were also reported by a previous study^29^, which might reflect complex biology of OAS genes for innate immune response. In a recent transcription-wide association study from the GenOMICC program^22^, genetically-predicted high expression of OAS3 in lungs and whole blood were associated with higher risk of becoming critically ill COVID-19 patients. Although further studies to assess the roles of OAS genes specific to SARS-CoV-2 are needed, it is likely that OAS1 is the main driver of the protective effect of Neanderthal haplotype for COVID-19 outcomes given prior functional studies demonstrating the antiviral effect of OAS genes^29^.

*IL10RB* encodes for the beta subunit of the IL10 receptor (a type III interferon receptor), and is part of a cluster of immunologically important genes including *IFNAR1* and *IFNAR2*, both recently implicated in severe COVID-19 pathophysiology.^58^ *IFNAR1* and 2 encode the interferon alpha/beta receptor subunits 1 and 2, respectively. Interestingly, while there exists a *cis*-pQTL strongly associated with IFNAR1 levels, it was not associated with any of the COVID-19 outcomes (P ∼ 0.5). Further, *IFNAR1* had no *trans*-pQTLs identified, which means that the *IL10RB cis*-pQTL does not likely reflect IFNAR1 levels. However, since IFNAR2 was not measured in any proteomic studies, we could not test the effect of its circulating levels on COVID-19 outcomes. IL10RB mediates IL10 anti-inflammatory activity through its downstream inhibitory effect on many well-known pro-inflammatory cytokines such as janus kinases and STAT1.^59^ While overexpression of IL10 has been involved in the persistence of multiple chronic bacterial infections such as tuberculosis,^60^ its role remains poorly understood in acute infections. In sepsis, a disease state characterized by high levels of cytokine activity and a rise in multiple biomarkers associated with inflammation, there is also a well-established increase in anti-inflammatory IL10 production by leukocytes, especially in the early stage of the disease.^61^ Most importantly, while in a normal physiological state, IL10 is usually only produced at a low level by neutrophils, monocytes and macrophages, its production is strongly upregulated by IL4, itself upregulated by lipopolysaccharides (LPS) when they bind LBPs.^62,63^ Interestingly, while the LBP gene did not pass FDR correction, it was still one of the most significant protein in our MR *cis-*pQTL analysis (**Table S1, S3**). While LPS’s are well-known for their role in triggering gram-negative bacterial sepsis, their role in other acute infections and respiratory diseases is likely broader, and involves complex sequences of cytokine signaling.^64–67^ Nevertheless, as our MR studies showed that IL10RB protein level affected COVID-19 outcome with a concordant effect direction, and given the known role of overt inflammation in COVID-19 morbidity, this pathway likely deserves more investigation.

This study has limitations. First, we used MR to test the effect of circulating protein levels measured in a non-infected state. This is because the effect of the *cis*-pQTLs upon circulating proteins was estimated in individuals who had not been exposed to SARS-CoV-2. Once a person contracts SARS-CoV-2 infection, levels of circulating proteins could be altered and this may be especially relevant for cytokines such as IL10 (which binds to IL10RB), whose levels may reflect host response to the viral infection and OAS1, whose levels are increased by activation of interferon pathway, as we observed in our case-control study (**Figures S4, S6, S9**). Thus, the MR results presented in this paper should be interpreted as an estimation of the effect of circulating protein levels, when measured in the non-infected state. On-going studies will help to clarify if the same *cis-*pQTLs influence circulating protein levels during infection. Second, this type of study suffers a high false-negative rate. Our goal was not to identify *every* circulating protein influencing COVID-19 outcomes, but rather to provide evidence for few proteins with strong *cis*-pQTLs since these proteins are more likely to be robust to the assumptions of MR studies. Future large-scale proteomic studies with more circulating proteins properly assayed should help to overcome these limitations. Third, most MR studies assume a linear relationship between the exposure and the outcome. Thus, our findings would not identify proteins whose effect upon COVID-19 outcomes has a clear threshold effect. Finally, we could not completely exclude the possibility that measurement of OAS1 levels may be influenced by protein altering variants, however, such variants do not affect sQTL RNA-sequencing studies and the association between OAS1 levels and COVID-19 outcomes remained robust in such analyses.

In conclusion, we have used genetic determinants of circulating protein levels and COVID-19 outcomes obtained from large-scale studies and found compelling evidence that OAS1 has a protective effect on COVID-19 susceptibility and severity. Measuring OAS1 levels in a case-control study demonstrated that higher OAS1 levels in a non-infectious state were associated with reduced risk of COVID-19 outcomes. Interestingly, the available evidence suggests that the protective effect from OAS1 is likely due to the Neanderthal introgressed p46 OAS1 isoform. Known pharmacological agents that increase OAS1 levels could be explored for their effect on COVID-19 outcomes.

## Methods

### pQTL GWAS

We systematically identified pQTL associations from six large proteomic GWASs.^15–20^ Each of these studies undertook proteomic profiling using either SomaLogic^®^ technology, or O-link proximal extension assays.

### COVID GWAS and COVID-19 Outcomes

To assess the association of *cis*-pQTLs with COVID-19 outcomes, we used the largest COVID-19 meta-analytic GWAS to date from the COVID-19 Host Genetics Initiative^21^. For our study, we used three of these GWAS meta-analyses which included 25 cohorts of European ancestry and 1 cohort of admixed American ancestry, based on sample size and clinical relevance. These outcomes were very severe COVID-19, hospitalization due to COVID-19, and susceptibility to COVID-19 (named A2, B2, and C2, respectively in the COVID-19 Host Genetics Initiative).

Very severe COVID-19 cases were defined as hospitalized individuals with COVID-19 as the primary reason for hospital admission with laboratory confirmed SARS-CoV-2 infection (nucleic acid amplification tests or serology based), and death or respiratory support (invasive ventilation, continuous positive airway pressure, Bilevel Positive Airway Pressure, or continuous external negative pressure, high-flow nasal or face-mask oxygen). Simple supplementary oxygen (e.g. 2 liters/minute via nasal cannula) did not qualify for case status. Controls were all individuals in the participating cohorts who did not meet this case definition.

Hospitalized COVID-19 cases were defined as individuals hospitalized with laboratory confirmed SARS-CoV-2 infection (using the same microbiology methods as for the very severe phenotype), where hospitalization was due to COVID-19 related symptoms. Controls were all individuals in the participating cohorts who did not meet this case definition.

Susceptibility to COVID-19 cases were defined as individuals with laboratory confirmed SARS-CoV-2 infection, health record evidence of COVID-10 (international classification of disease coding or physician confirmation), or with self-reported infections (e.g. by questionnaire). Controls were all individuals in the participating cohorts who did not meet this case definition.

### Two-sample Mendelian randomization

We used two-sample MR analyses to screen and test potential circulating proteins for their role influencing COVID-19 outcomes. In two-sample MR, the effect of SNPs on the exposure and outcome are taken from separate GWASs. This method often improves statistical power, because it allows for larger sample sizes for the exposure and outcome GWAS.^68^

Exposure definitions: We conducted MR using six large proteomic GWAS studies.^15–20^ Circulating proteins from Sun *et al*, Emilsson *et al* and Pietzner *et al* were measured on the Somalogic platform, Suhre *et al*, Yao *et al* and Folkersen *et al* used protein measurements on the O-link platform. We selected proteins with only *cis-pQTL*s to test their effects on COVID-19 outcomes, because they are less likely to be affected by potential horizontal pleiotropy. The *cis-pQTL*s were defined as the genome-wide significant SNPs (P < 5 × 10^−8^) with the lowest P value within 1 Mb of the transcription start site (TSS) of the gene encoding the measured protein.^9^ For proteins from Emilsson *et al*, Pietzner *et al*, Suhre *et al*, Yao *et al* and Folkersen *et al*, we used the sentinel *cis-*pQTL per protein per study as this was the data available. For proteins from Sun *et al*, we used PLINK and 1000 genome European population references (1KG EUR) to clump and select LD-independent *cis-*pQTL (R2<0.001, distance 1000 kb) with the lowest P-value from reported summary statistics for each SOMAmer^®^ bound proteins. We included the same proteins represented by different *cis*-pQTLs from different studies in order to cross examine the findings. For *cis-*pQTLs that were not present in the COVID-19 GWAS, SNPs with LD R^2^>0.8 and with minor allele frequency (MAF) < 0.42 were selected as proxies, MAF > 0.3 was used for allelic alignment for proxy SNPs. *cis-*pQTLs with palindromic effects and with minor allele frequency (MAF) > 0.42 were removed prior to MR to prevent allele-mismatches. Benjamini & Hochberg correction was used to control for the total number of proteins tested using MR. We recognize that this is an overly conservative correction, given the non-independence of the circulating proteins, but such stringency should reduce false positive associations. MR analyses were performed using the TwoSampleMR package in R.^69^ For proteins with a single (sentinel) *cis*-pQTL, we used the Wald ratio to estimate the effect of each circulating protein on each of the three COVID-19 outcomes. For any proteins/SOMAmer^®^ reagents with multiple independent *cis*-pQTL, an inverse variance weighted (IVW) method was used to meta-analyze their combined effects. After harmonizing the *cis-*pQTLs of proteins with COVID-19 GWAS, a total of 566 SOMAmer^®^ reagents (529 proteins, 565 directly matched IVs and 26 proxies) from Sun *et al*, 760 proteins (747 directly matched IVs and 11 proxies) from Emilsson *et al*, 91 proteins (90 directly matched IVs and 2 proxies) from Pietzner *et al*, 74 proteins (72 directly matched IVs) from Suhre *et al*, 24 proteins (24 directly matched IVs) from Yao *et al* and 13 proteins (13 directly matched IVs) from Folkersen *et al* were used as instruments for the MR analyses across the three COVID-19 outcomes (**Table S11-12**).^15–20^

### Pleiotropy assessments

A common pitfall of MR is horizontal pleiotropy, which occurs when the genetic variant affects the outcome via pathways independent of circulating proteins. The use of circulating protein *cis-pQTL*s greatly reduces the possibility of pleiotropy, for reasons described above. We also searched in the PhenoScanner database, a large catalogue of observed SNP-outcome relationships involving > 5,000 GWAS done to date to assess potentially pleiotropic effects of the *cis*-pQTLs of MR prioritized proteins, by testing the association of *cis*-pQTLs with other circulating proteins (i.e. if they were *trans*-pQTLs to other proteins or traits). For *cis*-pQTLs of MR prioritized proteins, if they were measured on SomaLogic^®^ platform, we assessed the possibility of potential aptamer-binding effects (where the presence of protein altering variants may affect protein measurements). We also checked if *cis*-pQTLs of MR prioritized proteins had significantly heterogeneous associations across COVID-19 populations in each COVID-19 outcome GWAS.

### Colocalization analysis

Finally, we tested colocalization of the genetic signal for the circulating protein and each of the three COVID-19 outcomes using colocalization analyses, which assess potential confounding by LD. Specifically, for each of these MR significant proteins with genome-wide summary data available, for the proteomic GWASs, a stringent Bayesian analysis was implemented in *coloc* R package to analyze all variants in 1MB genomic locus centered on the *cis*-pQTL. Colocalizations with posterior probability for hypothesis 4 (PP4, that there is an association for both protein level and COVID-19 outcomes and they are driven by the same causal variant) > 0.5 were considered likely to colocalize (which means the highest posterior probability for all 5 *coloc* hypotheses), and PP4 > 0.8 was considered to be highly likely to colocalize.

### sQTL and eQTL MR and colocalization studies for OAS genes

We performed MR and colocalization analysis using GTEx project v8^33^ GWAS summary data to understand the effects of expression and alternative splicing of *OAS* genes in whole blood. The genetic instruments were conditionally independent (R^2^ < 0.001) sQTL and eQTL SNPs for *OAS1*, eQTL for *OAS2* and *OAS3* identified by using stepwise regression in GTEx^33^. The sQTL SNP for *OAS1* (rs10774671), was originally identified for the normalized read counts of LeafCutter^31^ cluster of the last intron of p46 isoform (chr12:112,917,700-112,919,389 GRCh38) in GTEx^32^, and was used to estimate the effect of p46 isoform. Colocalization analysis was performed using GWAS summary from GTEx by restricting the regions within 1 Mb of rs4767027.

### Measurement of plasma OAS1 protein levels associated with COVID-19 outcomes in BQC19

BQC19 is a Québec-wide initiative to enable research into the causes and consequences of COVID-19 disease. For this analysis, we used results from patients with available proteomic data from SomaLogic^®^ assay (**Supplementary Data**). The patients were recruited at the Jewish General Hospital (JGH) and Centre hospitalier de l’Université de Montréal (CHUM) in Montréal, Québec, Canada.

COVID-19 case – control status was defined to be consistent with the GWAS study from COVID-19 HGI, from which the MR results were derived. Namely, we tested the association of OAS1 protein levels with the three different COVID-19 outcome definitions both in samples procured from non-infected samples and from samples during the acute phase of the infection. The three outcomes were: 1) Very severe COVID-19—defined as hospitalized individuals with laboratory confirmed SARS-CoV-2 infection (nucleic acid amplification tests or serology based), and death or respiratory support (invasive ventilation, continuous positive airway pressure, Bilevel Positive Airway Pressure, or continuous external negative pressure, high flow nasal or face-mask oxygen). Controls were all individuals who did not meet this case definition; 2) Hospitalized COVID-19 cases—defined as individuals hospitalized with laboratory confirmed SARS-CoV-2 infection. Controls were all who did not meet this case definition; 3) Susceptibility to COVID-19—cases were defined as individuals with laboratory confirmed SARS-CoV-2 infection, and controls were all individuals who underwent PCR testing for SARS-CoV-2, but were negative. The date of symptom onset for COVID-19 patients was collected from patients’ charts or estimated from their first positive COVID-19 tests if missing. Case inclusion criteria was not exclusive, which means that some individuals who were cases in the susceptibility analyses were also included in the hospitalization and very severe COVID-19 if they met case definitions.

Among SARS-CoV-2 positive participants, we defined samples procured from participants during the infectious state as those sampled within 14 days (including the 14^th^ day) from the first date of symptoms^70^. For individuals with more than one sample within 14 days of symptom onset, the earliest sample was used. We defined samples procured from patients who were non-infectious as samples from SARS-CoV-2 positive patients taken at least 31 days after symptom onset or from SARS-CoV-2 negative individuals. We selected 31 days, as this is the upper limit of the intra-quartile range of the duration of SARS-CoV-2 positivity in a recent systematic review and coincided with the first scheduled outpatient follow-up blood test in the BQC19^71^. For individuals with more than one sample at least 31 days of symptom onset, the latest sample was used. Protein levels in citrated (ACD) plasma samples were measured using the SomaScan®? assay [SomaLogic Inc.]. Details regarding SOMAmer QC are included in **Supplementary Data**.

1039 samples from 399 SARS-CoV-2 positive patients and 105 SARS-CoV-2 negative patients of mainly European descent underwent SomaScan^®^ assays, which included 5,284 SOMAmer reagents, targeting 4,742 proteins. A total of 125 individuals were recruited from CHUM and 279 individuals were recruited from the JGH. Individuals had blood sampling done at up to five different time points (200 individuals had one measurement, 113 individuals had two measurements, 152 individuals had three measurements, 38 individuals had four measurements and 1 individual had five measurements). Days from symptom onset were calculated for each sample based on the date of symptom and blood draw date. Sample processing time (in hours) for each sample was also calculated measure the duration of time from sample collection to processing to account for the changes in the amount of protein released from cell lysis due to sample handling time.

Sample QC was performed to remove outliers with long sample processing time and high OAS1 levels. OAS1 level was measured by one SOMAmer reagent (OAS1.10361.25). Within each group, normalized OAS1 levels were natural log transformed, adjusted for sample processing time and the residuals were further standardized. Logistic regression was performed to test the association standardized OAS1 level with the three COVID-19 outcomes including age, sex, age^2^, center of recruitment and plates as covariates.

## Data Availability

Data from proteomics studies and GTEx consortium are available from the referenced peer-reviewed studies or their corresponding authors, as applicable. Summary statistics for the COVID-19 outcomes are publicly available for download on the COVID-19 HGI website (www.covid19hg.org). Applicants are invited to apply for access to BQC19 data from the JGH hospital (https://www.mcgill.ca/genepi/mcg-covid-19-biobank) and/or the BQC19 (bqc19.ca).

## Acknowledgements

We thank Dr. Luis Barriero for his comments on the introgressed Neanderthal p46 isoform, and Dr. Nicolas Chomont for the discussion on the clinical phenotype of BQC19. We would like to also thank the MI4 and the MUHC Foundation that contributed for the SomaLogic^®^ panel.

## Ethics declarations

All cohorts contributing cohorts to COVID-19 HGI received ethics approval from their respective research ethics review boards. The Biobanque Quebecoise de la COVID-19 (BQC19) received ethical approval from the IRB of JGH and the CHUM.

## Author contributions

Conception and design: SZ, GBL and JBR. Data analyses: SZ and TN. Data acquisition: TN, GBL, DM, DEK, JA, MA, LL, EBR, DH, NK, ZA, NR, MB, LP, CG, XX, CT, BV, OA, TA, NA, MC, MD, VF, DEK and JBR. Interpretation of data: SZ, GBL, TN, MP, YC, DEK, VF and JBR. Funding acquisition: DM, VM, VF, JBR. Methodology: SZ, KZ, CMTG and JBR. Project administration: DM, VF and JBR. Validation: SZ, TN, MP, NK, MP, JN, ET, CL, DEK and JBR. Visualization: SZ, TN and VF. Writing-original draft: SZ, GBL, TN and JBR. Writing-review & editing: SZ, GBL, TN, MP, HZ, VM, MP, RF, ML, MH, CP, DEK and JBR. All authors were involved in preparation of the further draft of the manuscript and revising it critically for content. All authors gave final approval of the version to be published. The corresponding author attests that all listed authors meet authorship criteria and that no others meeting the criteria have been omitted.

## Notes

**Funding:** The Richards research group is supported by the Canadian Institutes of Health Research (CIHR: 365825; 409511), the Lady Davis Institute of the Jewish General Hospital, the Canadian Foundation for Innovation (CFI), the NIH Foundation, Cancer Research UK, Genome Québec, the Public Health Agency of Canada, the **McGill** Interdisciplinary Initiative in Infection and Immunity and the Fonds de Recherche Québec Santé (FRQS). SZ is supported by a CIHR fellowship and a FRQS fellowship. GBL is supported by the a CIHR scholarship, and a joint FRQS and Québec Ministry of Health and Social Services scholarship. TN is supported by Research Fellowships of Japan Society for the Promotion of Science (JSPS) for Young Scientists. JBR is supported by a FRQS Clinical Research Scholarship. Support from Calcul Québec and Compute Canada is acknowledged. TwinsUK is funded by the Welcome Trust, Medical Research Council, European Union, the National Institute for Health Research (NIHR)-funded BioResource, Clinical Research Facility and Biomedical Research Centre based at Guy’s and St Thomas’ NHS Foundation Trust in partnership with King’s College London. BQC-19 is funded by FRQS, Genome Québec and the Public Health Agency of Canada. These funding agencies had no role in the design, implementation or interpretation of this study. VM is supported by a Canada Excellence Research Chair. The Kaufmann lab COVID-19 work is supported by the CIHR/CITF (VR2-173203), AmFAR (110068-68-RGCV) the CFI and FRQS. RF is supported by Swedish Research Council (2014-02569 and 2014-07606). MH is supported by a SciLifeLab/KAW national COVID-19 research program project grant (KAW 2020.0182).

### Funding Statement

The Richards research group is supported by the Canadian Institutes of Health Research (CIHR: 365825; 409511), the Lady Davis Institute of the Jewish General Hospital, the Canadian Foundation for Innovation (CFI), the NIH Foundation, Cancer Research UK, Genome Quebec, the Public Health Agency of Canada, the McGill Interdisciplinary Initiative in Infection and Immunity and the Fonds de Recherche Quebec Sante (FRQS). SZ is supported by a CIHR fellowship and a FRQS fellowship. GBL is supported by the a CIHR scholarship, and a joint FRQS and Quebec Ministry of Health and Social Services scholarship. TN is supported by Research Fellowships of Japan Society for the Promotion of Science (JSPS) for Young Scientists. JBR is supported by a FRQS Clinical Research Scholarship. Support from Calcul Quebec and Compute Canada is acknowledged. TwinsUK is funded by the Welcome Trust, Medical Research Council, European Union, the National Institute for Health Research (NIHR)-funded BioResource, Clinical Research Facility and Biomedical Research Centre based at Guy's and St Thomas' NHS Foundation Trust in partnership with King's College London. BQC-19 is funded by FRQS, Genome Quebec and the Public Health Agency of Canada. These funding agencies had no role in the design, implementation or interpretation of this study. VM is supported by a Canada Excellence Research Chair. The Kaufmann lab COVID-19 work is supported by the CIHR/CITF (VR2-173203), AmFAR (110068-68-RGCV) the CFI and FRQS. RF is supported by Swedish Research Council (2014-02569 and 2014-07606). MH is supported by a SciLifeLab/KAW national COVID-19 research program project grant (KAW 2020.0182).

