## Supplementary Data and Figures for "A Neanderthal OAS1 isoform Protects Against COVID-19 Susceptibility and Severity: Results from Mendelian Randomization and Case-Control Studies"

### Supplementary Methods and Results

#### SomaScan® Platform Quality Control

The SomaScan® assay uses single-stranded DNA aptamers ("SOMAmers"), which are designed to selectively bind to a particular protein target[1]. SOMAmer reagent binding is quantified by microarray, measuring abundance in Relative Fluorescent Units (RFU). The RFUs for each protein underwent four normalization processes including hybridization control, intraplate median signal normalization, plate scaling and calibration and median signal normalization to a reference generated from internal data across all samples. All normalizations were conducted by SomaLogic® and detailed in their Technical Note[2].

#### Sample Descriptions and QC of BQC-19 cohort:

Days from time of blood draw to symptom onset were calculated for each sample based on the symptom onset date and blood draw date (T1), and for COVID-19 negative individuals, T1 was set to 0. The distribution of days from symptom onset in patients and sample processing time with OAS1 level for all 1039 samples and samples included in the analyses after QC are shown in **Figure S5A**, **S5B, S5C, S5D,** respectively**.** Distribution of log transformed of OAS1 in samples in a non-infectious state and during infection showed increased mean OAS1 levels in samples during infection (**Figure S6**). We also observed a clear decline of OAS1 level in individual patients during the trajectory of infection and this decline was more pronounced in patients with very severe COVID-19 outcomes (**Figure S7**).

As a result, for each group, we removed samples that were outliers in the delay between sample collection and sample process (sample processing time > 50 hrs) or OAS1 level (log OAS1 level > 8). After sample QC, 308 patients with at least one sample collected during infection and 113 patients with at least one sample collected during a non-infectious state and 103 COVID-19 negative controls were included in the analyses.

We tested the association of measured OAS1 levels with age and sex, since these are two important determinants of COVID-19 outcomes. We found no association of OAS1 level with age or sex (**Figure S8**) in samples during a non-infectious state.

In samples of non-infectious state, OAS1 levels did not appear to correlate with sample processing time, however there was a small increase of OAS1 level with prolonged sample processing time in people with active COVID-19 infection (**Figure S9**).

### Supplementary Figures

#### Figure S1. Colocalization of the Genetic Determinants of ABO Plasma Protein Levels and COVID-19 Outcomes

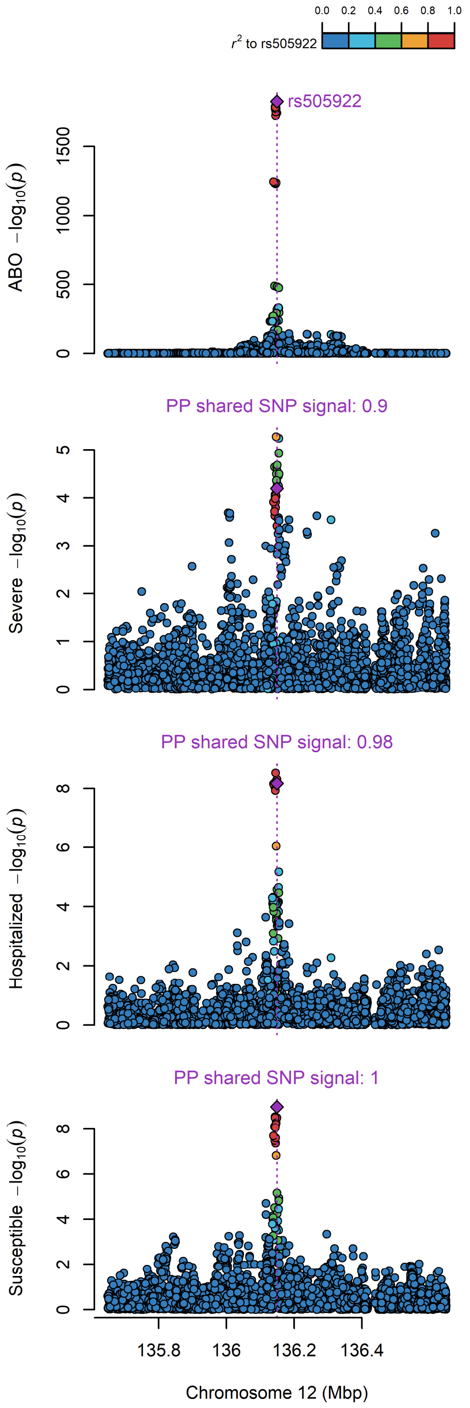


Color points represent the LD of SNPs within 1Mb region around the leading *cis*-pQTL of ABO rs505922. PP: posterior probability of sharing the same genetic signal to ABO level, estimated by *coloc*.

### Figure S2. Colocalization of the the expression QTL (eQTL), splicing QTL (sQTL) and protein QTL (pQTL) of OAS1 with COVID-19 outcomes

**(A) Locus plots for chr12 q24.13**

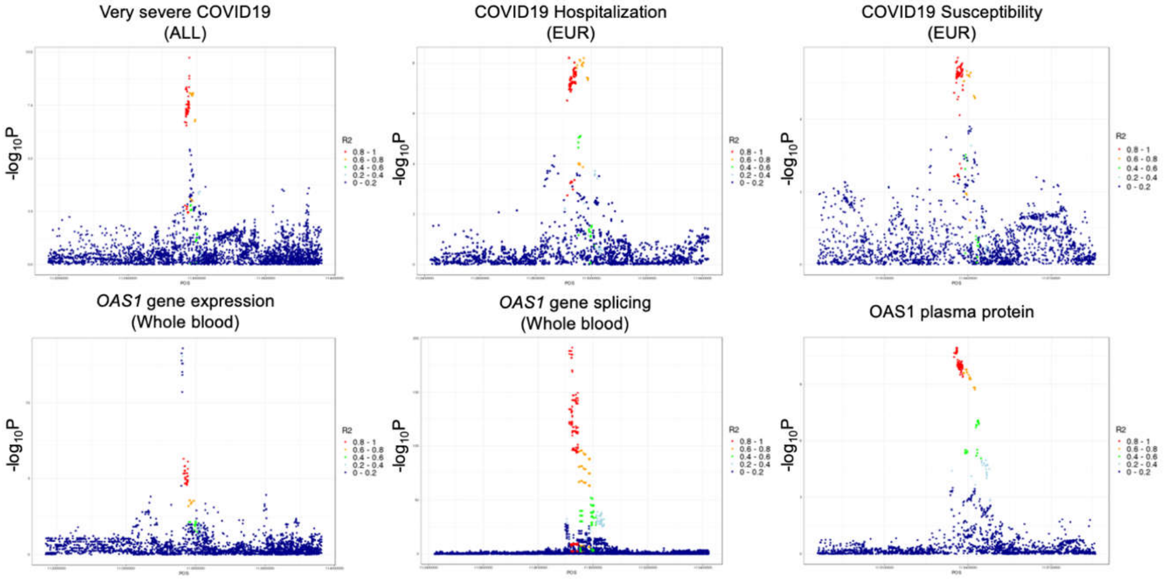


**(B) Colocalization analysis (PP4: posterior probability of shared causal signals)**

| | ***OAS1* Expression** | ***OAS1* Splicing** | **OAS1 Protein** | **Very severe COVID-19** | **COVID-19 Hospitalization** | **COVID-19 Susceptibility** |
|---|---|---|---|---|---|---|
| ***OAS1* Expression** | . | 6.4E-05 | 4.1E-05 | 1.2E-04 | 4.1E-05 | 4.9E-04 |
| ***OAS1* Splicing** | 6.4E-05 | . | 0.96 | 0.96 | 0.92 | 0.96 |
| **OAS1 Protein** | 4.1E-05 | 0.96 | . | 0.72 | 0.82 | 0.89 |

Legend: (A) R2 was calculated against the rs4767027 SNP in each COVID-19 GWAS using 503 European individuals in 1000 genome project[3]. (B) A table illustrating the pairwise colocalization results between OAS1 expression (eQTL), splicing (sQTL), protein (pQTL) and COVID-19 outcomes. Colocalization analysis was performed using GWAS summary from GTEx[4] by restricting the regions within 1 Mb of rs4767027.

**Figure S3. Colocalization of the expression QTL (eQTL) of *OAS3* and COVID-19 outcomes**

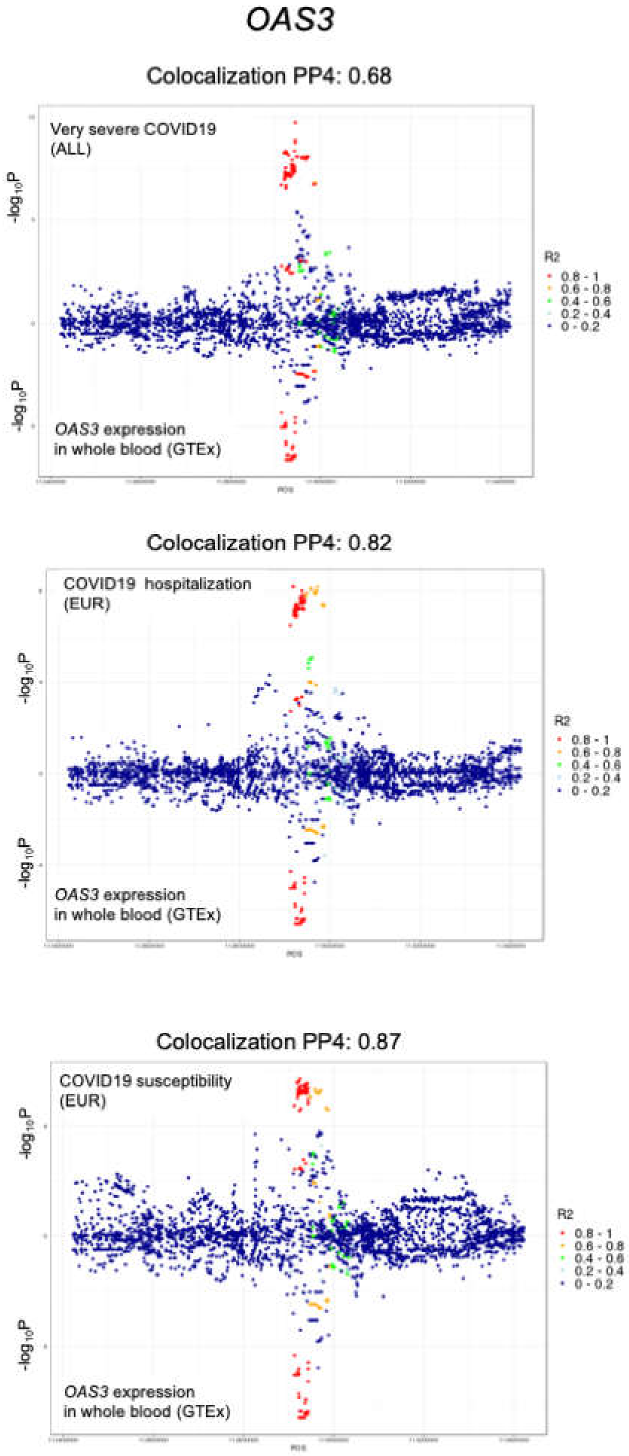


Legend: R2 was calculated against the SNP with the lowest p-value in each COVID-19 GWAS using 503 European individuals in 1000 genome project[3]. Colocalization analysis was performed using GWAS summary from GTEx[4] by restricting the regions within 1 Mb of rs4767027

**Figure S4. OAS1 Level Trajectory with Days Since Symptom Onset in different COVID-19 Outcomes**

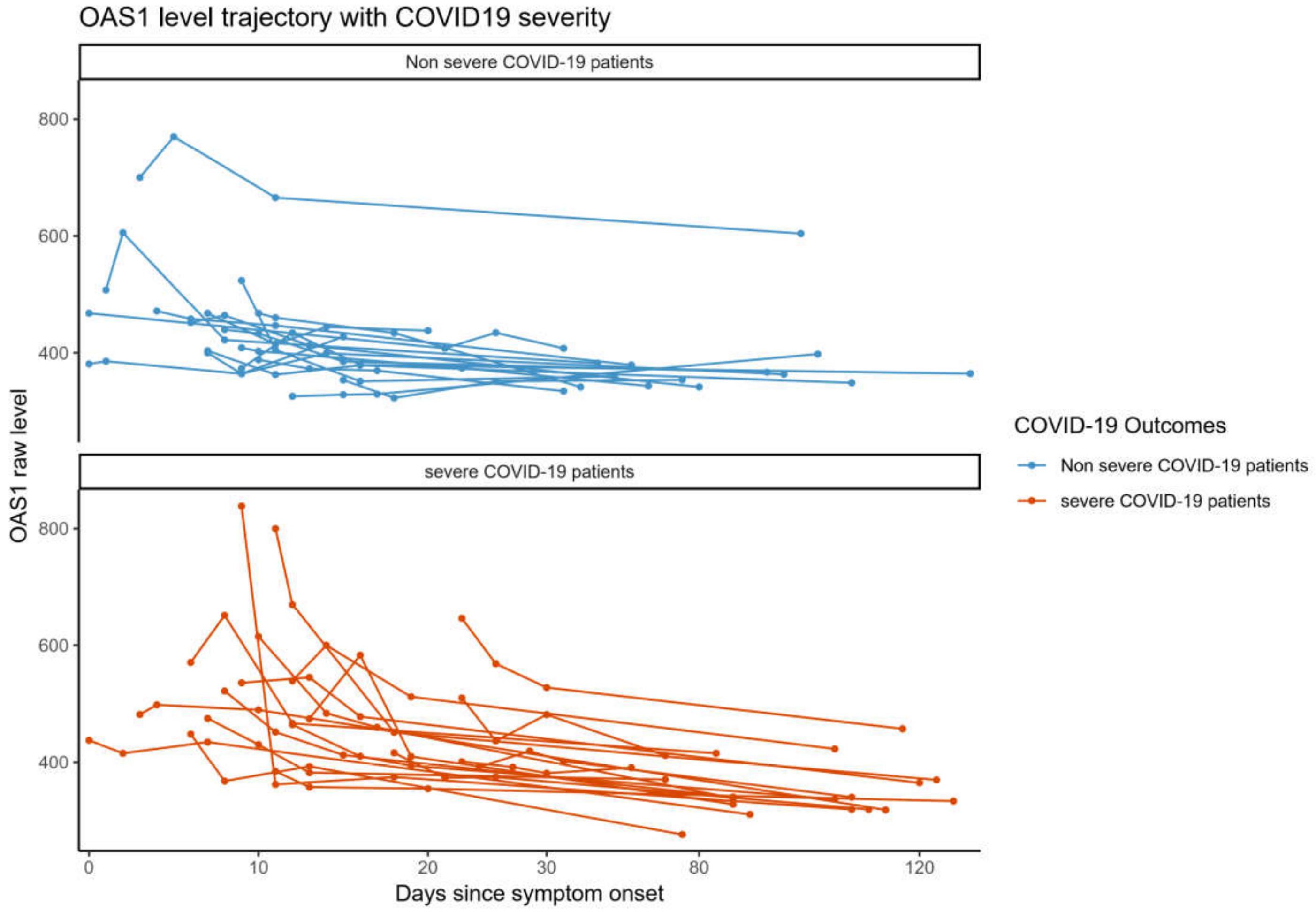


Trajectory plot showing the OAS1 level (in normalized RFUs) of 38 COVID-19 patients with their blood samples collected at four different time points, represented by days since symptom onset. Each line represents one patient. Blue shows COVID-19 patient with not severe outcome and red shows COVID-19 patient with severe outcome.

### Figure S5. Distribution of Days from Symptom Onset and Sample Processing Time in BQC-19 Samples

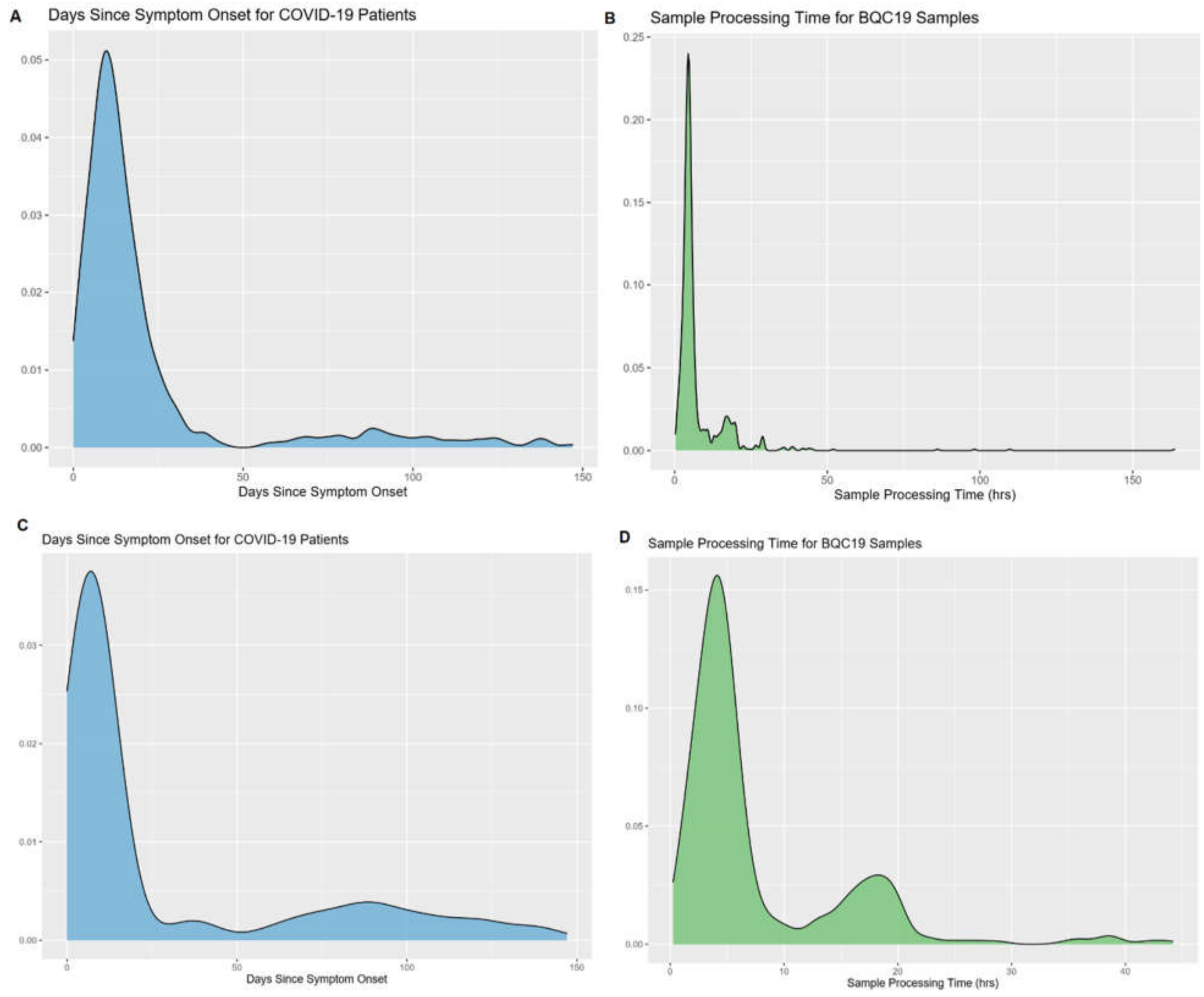


A: Distribution of Days from Symptom Onset in all 934 samples from COVID-19 patients with SomaScan® measurement.
B. Distribution of Sample Processing Time in all 1039 samples with SomaScan® measurement.
C. Distribution of Days from Symptom Onset in 421 samples from COVID-19 patients included in the analyses.
D. Distribution of Sample Processing Time in 627 samples included in the analyses.

**Figure S6. Distribution of log transformed of OAS1 in samples during non-infectious state and during infection**

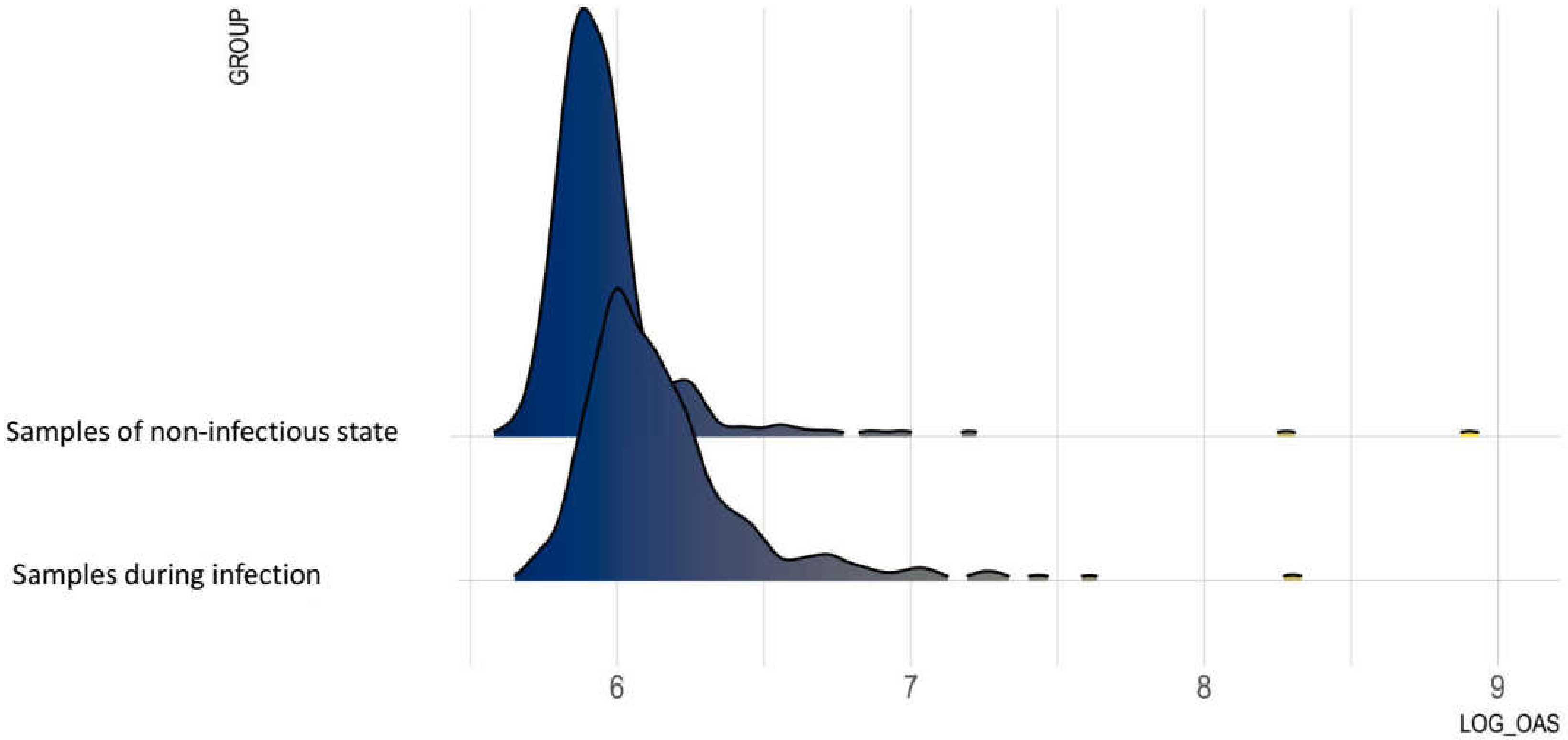


Figure shows after samples in each group before QC.

**Figure S7. OAS1 level association with Age and Sex in samples during non-infectious state**

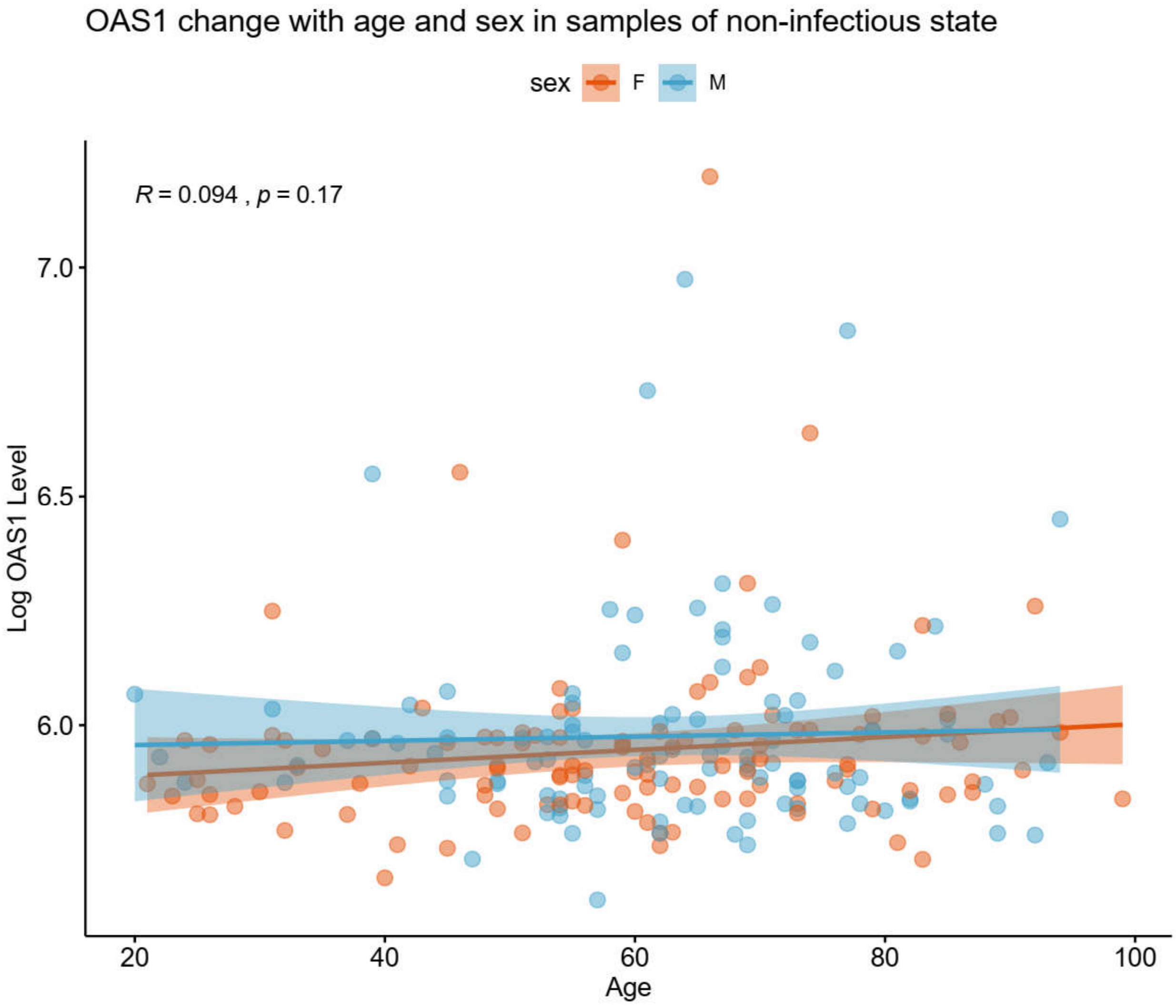


Figure shows after removal of outliers of OAS1 level and sample processing time.

**Figure S8. Violin plot of OAS1 Level Measured by SomaScan® Technology in BQC19 cohort in Groups of Different COVID-19 Outcomes, in samples taken from a Non-infectious State and During Infection.**

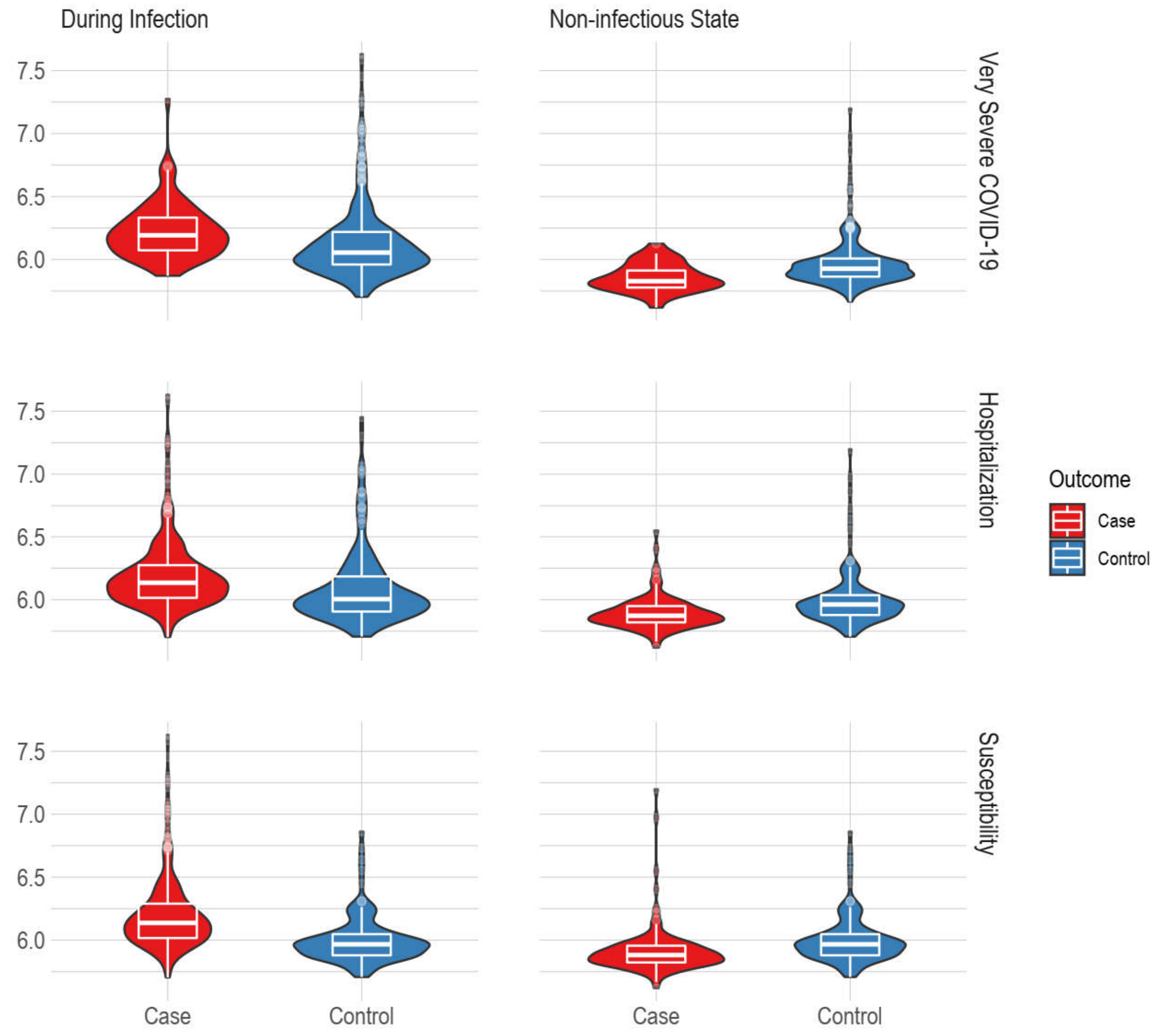


Y axis shows natural log transformed OAS1 level. Controls of each groups include COVID negative individuals.

**Figure S9. Correlation of log OAS1 level with Processing Time in Samples from a Non-infectious State and During Infection.**

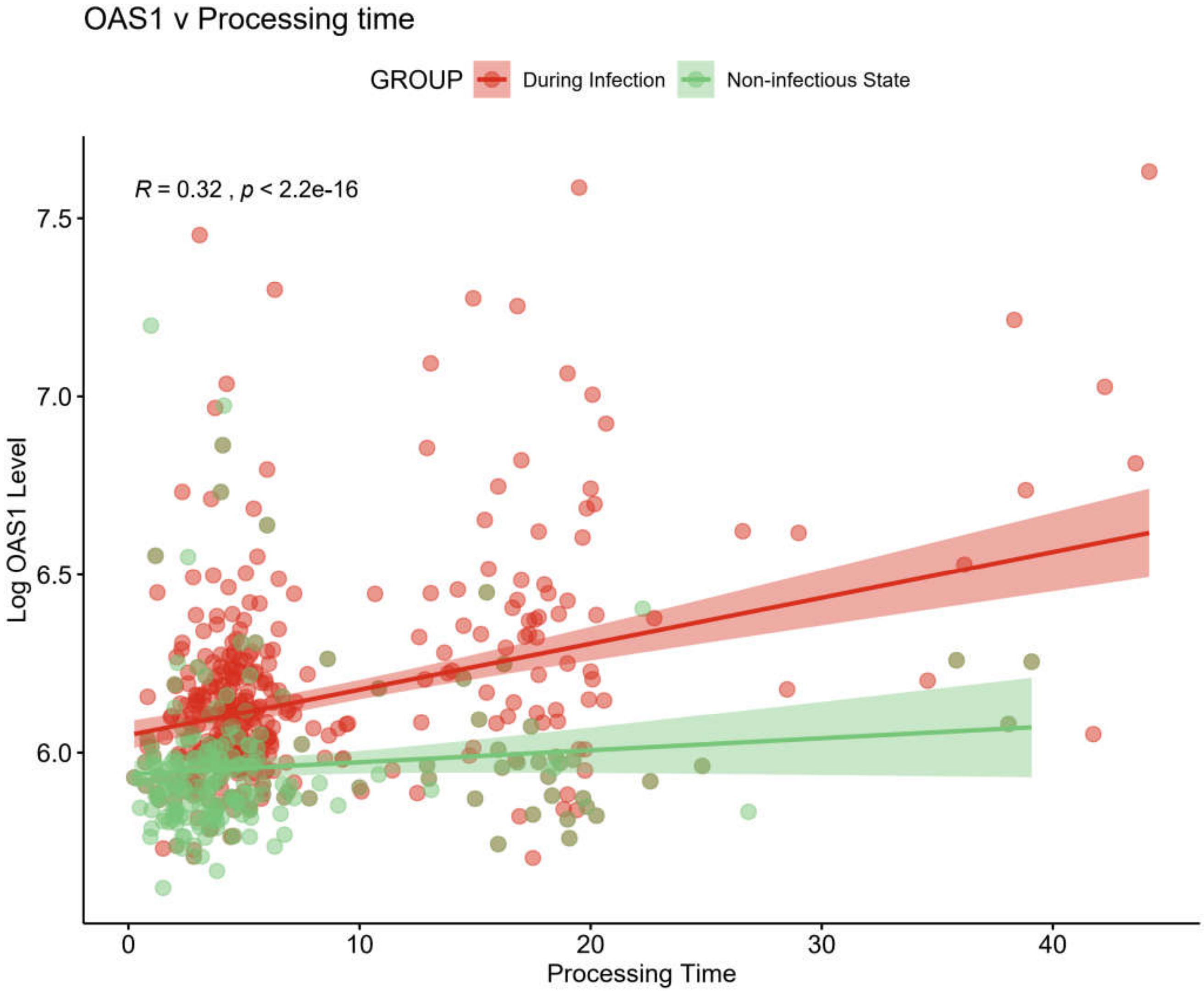


Figure shows after removal of outliers of OAS1 level and sample processing time.


---

ii
